## Appendix 1 Rapid Review of Literature for "Cross-sectional evaluation of an asynchronous Multiple Mini Interview (MMI) in selection to health professions training programmes with ten principles for fairness built-in"

Appendix A Rapid Review Key References in Health Professions Selection

Further details available on reasonable request


35. Ballejos, M. P., Oglesbee, S., Hettema, J., Sapien, R. An equivalence study of interview platform: Does videoconference technology impact medical school acceptance rates of different groups?. Advances in Health Sciences Education: Theory and Practice. 2018;23(3):601–610. https://doi.org/http://dx.doi.org/10.1007/s10459-018-9817-2 Page 16/21

36. Bird, SB, Hern, G, Blomkalns, A, Deiorio NM, Haywood Y, Hiller KM, et al. Innovation in Residency Selection: The AAMC Standardized Video Interview. Academic Medicine: Journal of the Association of American Medical Colleges. 2019;94(10):1489–1497. https://doi.org/https://dx.doi.org/10.1097/ACM.0000000000002705

37. Bowers, K., Comp, G., Kalnow, A., Casey, J., Fraser, W., Lloyd, C., Little, A. Are Standardized Video Interview Scores Predictive of Interview Performance? Western Journal of Emergency Medicine. 2018;S18

---

44. Egan, D. J., Husain, A., Bond, M. C., Caputo, W., Cygan, L., Van Dermark, J., et al. Standardized video interviews do not correlate to United States medical licensing examination step 1 and step 2 scores. Page 18/21 The Western Journal of Emergency Medicine. 2019;20(1):87–91. https://doi.org/http://dx.doi.org/10.5811/westjem.2018.11.39730

45. Edje, L., Miller, C., Kiefer, J., & Oram, D. Using Skype as an Alternative for Residency Selection Interviews. Journal of Graduate Medical Education. 2013;5(3):503–505. https://doi.org/10.4300/jgme-d-12-00152.1

46. Gallahue, F. E., Hiller, K. M., Bird, S. B., Calderone Haas, M. R., Deiorio, N. M., Hern, H. G., et al. The AAMC Standardized Video Interview: Reactions and Use by Residency Programs During the 2018 Application Cycle. Academic Medicine: Journal of the Association of American Medical Colleges. 2019;94(10):1506–1512. https://doi.org/https://dx.doi.org/10.1097/ACM.0000000000002714

47. Grova MM, Donohue SJ, Meyers MO, Kim HJ, Ollila DW. Direct Comparison of In-Person Versus Virtual Interviews for Complex General Surgical Oncology Fellowship in the COVID-19 Era. Ann Surg Oncol. 2021;28(4):1908-1915. doi:10.1245/s10434-020-09398-2

48. Hakes, E., Schnapp, B., Ritter, D., Kraut, A., Fallon, S., Brown, K., Westergaard, M. Communication and Professionalism: Comparing Standardized Video Interview Scores to Faculty Gestalt. Society for Academic Emergency Medicine. 2018;S276.

49. Hall, MM, Lewis, JJ, Joseph, JW, Ketterer, AR, Rosen, CL, Dubosh, NM. Standardized Video Interview Scores Correlate Poorly with Faculty and Patient Ratings. The Western Journal of Emergency Medicine. 2019;21(1):145–148. https://doi.org/https://dx.doi.org/10.5811/westjem.2019.11.44054

50. Healy, W. L., Bedair, H., & W.L., H. Videoconference Interviews for an Adult Reconstruction Fellowship: Lessons Learned. The Journal of Bone and Joint Surgery. American Volume. 2017;99(21):e114. https://doi.org/https://dx.doi.org/10.2106/JBJS.17.00322

51. Hopson, L. R., Dorfsman, M. L., Branzetti, J., Gisondi, M. A., Hart, D., Jordan, J., et al. Comparison of the Standardized Video Interview and Interview Assessments of Professionalism and Interpersonal Communication Skills in Emergency Medicine. AEM Education and Training. 2019;3(3):259–268. https://doi.org/https://dx.doi.org/10.1002/aet2.10346

52. Humbert, A., Pettit, K., Mugele, J., Turner, J., Morgan, Z., & Palmer, M. Correlation of the Standard Video Interview Score With an Established Application Review Process. Society for Academic Emergency Medicine. 2018;S98.

53. Huppert L A, Hsiao E C, Cho K C *et al.* Virtual interviews at graduate medical education training programs determining evidence-based best practices. *Acad Med* 2020. doi: 10.1097/ACM.0000000000003868.

---

54. Husain, A., Li, I., Ardolic, B., Bond, M. C., Shoenberger, J., Shah, K. H., et al. The Standardized Video Interview: How Does It Affect the Likelihood to Invite for a Residency Interview?. AEM Education and Training. 2019;3(3):226–232. https://doi.org/https://dx.doi.org/10.1002/aet2.10331

Inzana, K, Vanderstichel, R, Newman, S. Virtual Multiple Mini-Interviews for Veterinary Admissions. https://doi.org/10.3138/jvme-2020-0107 J Of Veterinary Medical Education

56. Joshi, A., Bloom, D. A., Spencer, A., Gaetke-Udager, K., & Cohan, R. H. Video Interviewing: A Review and Recommendations for Implementation in the Era of COVID-19 and Beyond. Academic radiology. 2020;27(9):1316–1322. https://doi.org/10.1016/j.acra.2020.05.020

57. Kok K, Chen L, Idris F, Mumin N, Ghani H, Zulkipli. Conducting multiple mini-interviews in the midst of COVID-19 pandemic. Korean J Med Educ. 2020 Dec; 32(4): 281–289. Published online 2020 Oct 28. doi: 10.3946/kjme.2020.175.

58. Krauss, W., Egan, D., Bond, M., Husain, A., White, M., Taylor, T., et al. Correlation Between Emergency Medicine Residency Applicant's Standardized Video Interview Scores and United States Medical Licensing Examination Results. Society for Academic Emergency Medicine. 2018;S83

Kyong-Jee Kim, Nam Young Lee & Bum Sun Kwon. Benefits and Feasibility of Using Videos to Assess Medical School Applicants' Empathetic Abilities in Multiple Mini Interviews. *Medical Science Educator* volume 31, pages175–181 (2021)C

60. Lewis, J., Hall, M., Joseph, J., Dubosh, N., Ullman, E. Standardized Video Interview Scores Do Not Correlate With Attending Evaluations. Society for Academic Emergency Medicine, 2018;S229.

61. M., S.R., E., L.M., B., M.M., A., F.M., D., J.C., E., … H., F. Initial Experience with a Virtual Platform for Advanced Gastrointestinal Minimally Invasive Surgery Fellowship Interviews. Journal of the American College of Surgeons. 2020;231(6):670–678. https://doi.org/http://dx.doi.org/10.1016/j.jamcollsurg.2020.08.768

62. McAteer, R., Sundaram, S., Harkisoon, S., & Miller, J. Videoconference Interviews: A Timely Primary Care Residency Selection Approach. Journal of Graduate Medical Education. 2020;12(6):737–744. https://doi.org/https://dx.doi.org/10.4300/JGME-D-20-00248.1

63. McHugh, M., Kulstad, C., Van Dermark, J., Bischof, J. Do Standardized or Traditional Interview Questions Correlate With the Standardized Video Interview? Society for Academic Emergency Medicine. 2019;S216

64. Molina, G., Mehtsun, W. T., Qadan, M., Hause, K. C., Raut, C. P., & Fairweather, M. Virtual Interviews for the Complex General Surgical Oncology Fellowship: The Dana-Farber/Partners Experience. Annals of Surgical Oncology. 2020;27(9):3103–3106. https://doi.org/https://dx.doi.org/10.1245/s10434-020- 08778-y

65. Nutter, A., La Rosa, M., Olson, G. Perception of Candidates and Faculty on Maternal Fetal Medicine Fellowship Videoconference Interviewing. Obstetrics and Gynecology. 2020;S75.

Nwora C, M.D.[a] Allred, D, M.D.[b] Verduzco-Gutierrez, M, M.D.[c] Mitigating Bias in Virtual Interviews for Applicants Who are Underrepresented in Medicine. Journal of the National Medical Association Volume 113, Issue 1, February 2021, Pages 74-76

67. Patel T, Bedi H, Deitte L, Lewis P *et al.* Brave New World: Challenges and Opportunities in the COVID-19 virtual interview season. *Acad Radiol* 2020; 27: 1456-1460.

68. Proost, K., Germeys, F., & Vanderstukken, A. Applicants' pre-test reactions towards video interviews: the role of expected chances to demonstrate potential and to use nonverbal cues.

European Journal of Work and Organizational Psychology. 2021;30(2):265-273. DOI: 10.1080/1359432X.2020.1817975

69. Sabesan V, Kapur N, Zemanek K, Levitt D, Vu T, Erp A. Implementation and evaluation of virtual multiple mini-interviews as a selection tool for entry into paediatric postgraduate training: A Queensland experience. Medical Teacher Published online: 30 Aug 2021.

https://doi.org/10.1080/0142159X.2021.1967906

70. Schnapp, B. H., Ritter, D., Kraut, A. S., Fallon, S., Westergaard, M. C. Assessing residency applicants' communication and professionalism: Standardized video interview scores compared to faculty gestalt. Western Journal of Emergency Medicine. 2019;20(1):132–137. https://doi.org/http://dx.di.org/10.5811/westjem.2018.10.39709

71. Shah SK, Arora, S, Skipper B, Kalishman S, Timm TC, Smith AY. Randomized evaluation of a web based interview process for urology resident selection. Journal of Urology. 2012;187(4):1380–1384. https://doi.org/http://dx.doi.org/10.1016/j.juro.2011.11.108

72. Singh, N D, DeMesa, DO, MPH, Pritzlaff, S, MD, Jung, M MD, MBA, Green, C MA, LMFT. Implementation of Virtual Multiple Mini-Interviews for Fellowship Recruitment . *Pain Medicine*, Volume 22, Issue 8, August 2021, Pages 1717–1721, https://doi.org/10.1093/pm/pnab141

73. Sripad, A. Videoconference Interviews for Female Pelvic Medicine and Reconstructive Surgery Fellowship During a Pandemic: The Candidate Experience. Female Pelvic Medicine & Reconstructive Surgery. 2020;S181.

74. Staicu, M., Hamby, C., Wychowski, M., Reiss, B. Facetime faceoff: evaluation of video conferencing as a novel pre-interview screen for a PGY-1 pharmacy residency. 2015;E182.

75. Temple, M., Lagzdins, M. Streamlining the residency interview process using Web-based teleconferencing. American Journal of Health-System Pharmacy. 2014;71:697-701. Doi: 10.2145/ajhp130406

76. Tiller D, O'Mara D, Rothnie I, Dunn S, Lee L, Roberts C. Internet-based multiple mini-interviews for candidate selection for graduate entry programmes. Med Educ. 2013;47(8):801-810. doi:10.1111/medu.12224

77. Turpin C, Steele K, Matuk-Villazon O, Rowland, K, Dayton, Horn, K. Rapid Transition to a Virtual Multiple Mini-Interview Admissions Process: A New Medical School's Experience During the COVID-19 Pandemic. Academic Medicine: August 2021 - Volume 96 - Issue 8 - p 1152-1155. doi: 10.1097/ACM.0000000000004179.

78. Ungtrakul T, Lamlertthon W, Boonchoo B, ,Auewarakul C. Virtual Multiple Mini-Interview during the COVID-19 Pandemic. Medical Education Adaptation. **https://doi.org/10.1111/medu.14207**

79. Vadi, M. G., Malkin, M. R., Lenart, J., Stier, G. R., Gatling, J. W., & Applegate, R. L. Comparison of web-based and face-to-face interviews for application to an anaesthesiology training program: a

pilot study. International Journal of Medical Education. 2016;7:102–108. https://doi.org/10.5116/ijme.56e5.491a

80. Vining CC, Eng OS, Hogg ME, et al. Virtual Surgical Fellowship Recruitment During COVID-19 and Its Implications for Resident/Fellow Recruitment in the Future. Ann Surg Oncol. 2020;27(Suppl 3):911- 915. doi:10.1245/s10434-020-08623-2

81. Willis, J., Surles, T., Silverberg, M., Kendall, S., LoCascio, H., Gernsheimer, J., Schechter, J., Regan, A., Smith, T. Are Standardized Video Interview Scores Predictive of Interview Performance? Western Journal of Emergency Medicine. 2018;S5-S6.

82. Winfield-Dial, A., Chhabra, N., Schindlbeck, M., & Bowman, S. Demographic Differences Between High and Low Scorers on the Standardized Video Interview. Western Journal of Emergency Medicine. 2018;S48. https://escholarship.org/uc/item/68c0x4pf

83. Winfield-Dial, A., Chhabra, N., Schindlbeck, M., & Bowman, S. Applicant Attitudes Towards the Standardized Video Interview - An Interim Analysis. Western Journal of Emergency Medicine. 2018;S4. https://escholarship.org/uc/item/5xp7f587 Page 17/21

84. Yolanda,S, Wisnu, W, Wahjudi, J and Findyartini A. Adaptation of internet-based multiple mini-interviews in a limited-resource medical school during the coronavirus disease 2019 pandemic. Korean J Med Educ. 2020 Dec; 32(4): 281–289. doi: 10.3946/kjme.2020.175
