## Appendix 2 for "Cross-sectional evaluation of an asynchronous Multiple Mini Interview (MMI) in selection to health professions training programmes with ten principles for fairness built-in"

Appendix 1. Results of a parallel analysis using Pearson correlations on the response data to the SAMMI MMI.

| Number of postulated factors | Mean of real data % of variance | Mean of random % of variance | 95 percentile of random % of variance |
|---|---|---|---|
| 1 | 29.2* | 2.9 | 3.1 |
| 2 | 11.1* | 2.8 | 3.0 |
| 3 | 9.5* | 2.8 | 2.9 |
| 4 | 8.3* | 2.7 | 2.8 |
| 5 | 7.6* | 2.6 | 2.7 |
| 6 | 7.4* | 2.6 | 2.7 |
| 7 | 6.2* | 2.5 | 2.6 |
| 8 | 1.1 | 2.5 | 2.5 |
| 9 | 1.0 | 2.4 | 2.5 |

*Percentage of variance in the responses explained by the number of factors in the real data exceeded that for the random data.
