## Appendix 3 for "Cross-sectional evaluation of an asynchronous Multiple Mini Interview (MMI) in selection to health professions training programmes with ten principles for fairness built-in"

Appendix 2. Estimated hierarchical confirmatory factor analytic (CFA) model for the SAMMI responses, with a general factor (g) and seven specific factors (f1 to f7), the latter representing the seven questions in the interview. Standardised factor loading are shown.

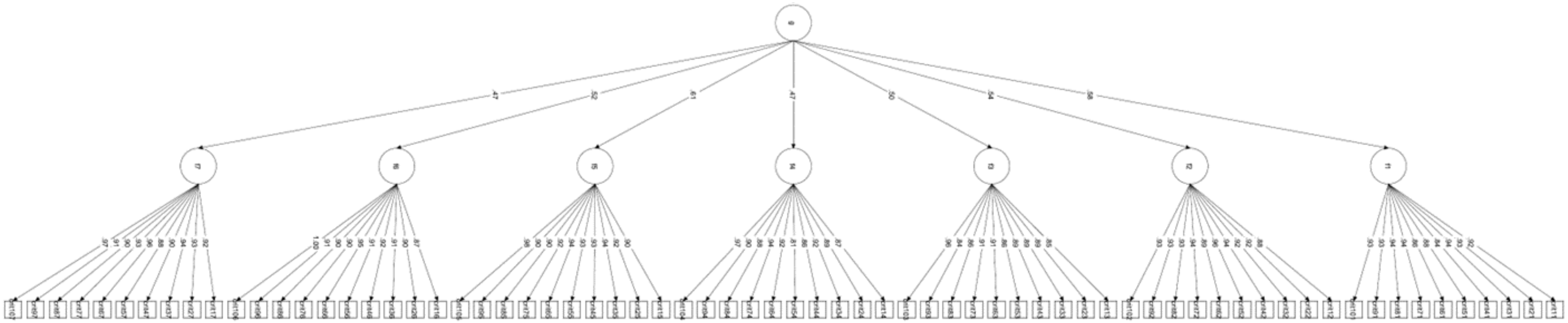
